# Transpulmonary proteome gradients identify pathways involved in the development of pulmonary vascular disease in heart failure

**DOI:** 10.1101/2025.02.01.25321512

**Authors:** Vojtech Melenovsky, Petr Jarolim, Eva Kutilkova, Dominik Jenca, Jana Binova, Hikmet Al-Hiti, Janka Franekova, Sona Kikerlova, Marketa Adamova, Matus Miklovic, Barry A. Borlaug

## Abstract

**Background and Aims:** Some, but not all, patients with heart failure (HF) develop pulmonary vascular disease (PVD), which contributes to poor prognosis. Mechanisms leading to PVD in HF are poorly understood. Unbiased analysis of transpulmonary gradients may identify mediators of PVD by finding proteins consumed or elaborated across the lungs.

**Methods:** 21 controls and 160 HFrEF patients underwent pulmonary artery (PA) catheterization with blood sampling from postcapillary (wedged balloon) and precapillary (unwedged) position to obtain transpulmonary gradients. The samples from controls and HF from the highest (Q4, n=40) and lowest quartile (Q1, n=40) of pulmonary vascular resistance (PVR) were analyzed using proteomic proximity extension assay (Olink) of 275 proteins. Venous blood concentrations or transpulmonary gradients were analyzed to identify biomarkers or mediators of PVD.

**Results:** Comparison of Q1 and Q4 of PVR identified Surfactant protein-D (PSP-D) as a marker of PVD. Examination of gradients across the lungs in high PVR HF revealed significant uptake of 18 mediators, mostly associated with inflammation (chemokines, oncostatin-M, MMP9) or with TGF/activin pathway (GDF2/BMP9), and release of 5 mediators, notably IL-6 and IL-33. In contrast, these protein gradients were negligible in controls and low PVR HF patients.

**Conclusions:** The lungs of patients with HF and high PVR, display abnormal uptake and release of proinflammatory cytokines from IL6/gp130 family (IL6, IL33, Oncostatin-M), along with increased transpulmonary uptake of GDF2/BMP9. The study shows that proteins orchestrating inflammation or pulmonary vessel remodeling in group 1 PH, are also operating in patients with PVD due to HF.

**Structured graphical abstract:** *Key question:* Can transpulmonary gradients of regulatory peptides, measured by unbiased proteomic assay, provide a hint on mechanisms of increased pulmonary vascular resistance (PVR) in patients with heart failure (HF)?

*Key findings:* In striking contrast to Controls (n=21) and HF with low PVR (n=40), the lungs of patients with HF and high PVR (n=40) display abnormal release of proinflammatory cytokines (IL6, IL33), along with increased transpulmonary uptake of chemokines, BNP, Oncostatin M and GDF2/BMP9, a key ligand of BMPRII receptor.

*Take home message:* The study shows that proteins orchestrating inflammation or pulmonary vessel remodeling in group 1 pulmonary hypertension, are also operating in patients with pulmonary vascular disease due to HF. 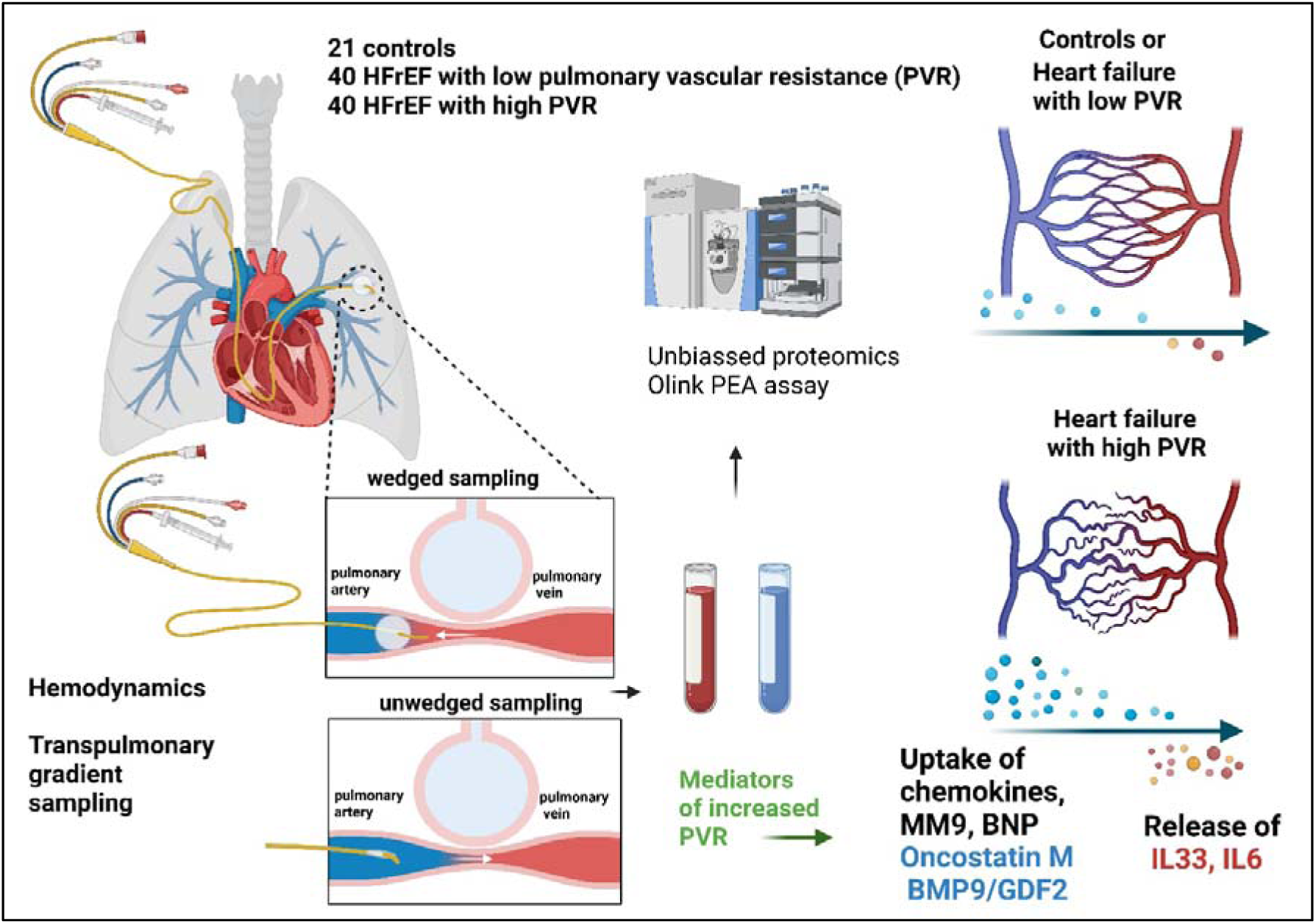

## Introduction

Some, but not all, patients with left-sided heart failure (HF) develop pulmonary vascular disease (PVD), which increases right ventricular afterload and promotes transition to biventricular HF, with poor prognosis^1–3^. PVD is defined hemodynamically by an increase in pulmonary vascular resistance (PVR). Histopathologic studies have shown that the morphological substrate of PVD is an intimal thickening in pulmonary venules and arterioles, with an absence of plexiform lesions typical for group 1 pulmonary hypertension (G1 PH)^3,4^. The mechanisms and signaling pathways involved in the development of PVD are poorly understood. This may explain in part why conventional pulmonary vasodilators, used for G1 PH, are not effective in patients with group 2 PH (G2 PH) due to underlying HF^3,5–7^.

PVD is more commonly present in patients with more advanced HF. Detailed analysis of venous blood samples may provide insight, but analyses of transpulmonary arteriovenous differences across an organ provides unique mechanistic insight to evaluate production and consumption of proteins or metabolites across the lung circulation^8,9^. In this approach, precapillary blood is sampled from pulmonary artery and postcapillary blood is obtained from wedged PA balloon catheter^10,11^. Here, we combined such sampling techniques with targeted proteomics by proximity extension assay,^12^ and compare patients with HF patients with and without PVD to healthy controls without HF or PVD^1^.

We reasoned, that net lung uptake of a regulatory protein is reflecting either cleavage, transendothelial transport or receptor-mediated removal, while net lung release reflect active secretion of the protein into pulmonary vessel tree^13^. If a transpulmonary gradient of a protein differs between patients with markedly different PVR, it implicates that the protein may be mechanistically linked to PVD. To test these hypotheses and to identify biomarkers or mediators of PVD in HF, we conducted a cross-sectional comparative study of controls and HFrEF patients with or without PVD, using top performing targeted proteomic technology^14^.

## Methods

### Patient cohorts and procedures

The study examined consecutive patients with chronic symptomatic HFrEF (left ventricular ejection fraction ≤40%) electively hospitalized for consideration of advanced therapies at the Institute for Clinical and Experimental Medicine (IKEM) in Prague, Czech Republic. Patients with acute ischemia, uncontrolled cardiac arrhythmia and hemodynamic instability requiring inotropes or mechanical circulatory support, reversible cardiac dysfunction, active malignancy, recent pulmonary embolism, endocrine disease, chronic or acute infection were excluded. Patients with hypervolemia on admission (based on clinical examination, increased body weight or measured central venous pressure) were enrolled into the study after diuresis and reaching normovolemia (central venous pressure <10[mmHg or return of body weight to normal with disappearance of clinical signs of congestion). The study complied with Declaration of Helsinki and the local ethics committee approved the protocol, the study is registered at ClinitralTrials.gov website (**NCT06331208**).

All subjects signed informed consent to participate in this research study. Patients with HF underwent history review, physical examination, echocardiography, electrocardiogram, Kansas City Cardiomyopathy Questionnaire (KCCQ) and morning (6-7 am) peripheral venous blood sampling in the post-absorptive state following an overnight fast. Control subjects were patients with normal echocardiography and hemodynamics, who underwent diagnostic RHC that ruled out cardiac disease (n=5) and subjects who underwent RHC as part of closure of patent foramen ovale (n=16). Right heart catheterization (RHC) was performed according to current recommendations using a 7[Fr balloon-tipped triple-lumen Swan–Ganz catheter (Braun AG, Germany) inserted via the right internal jugular vein (patients with HF) or femoral vein (controls referred for PFO closure). Hemodynamic zero was calibrated at mid-thoracic level. Pressure waveforms were recorded over several respiratory cycles and annotated by an invasive hemodynamic module (Mac-Lab, GE Healthcare, USA) as recommended^15^. Cardiac output (CO) was measured by thermodilution as an average of at least 3 measurements (of 5 in patient with atrial fibrillation) Pulmonary vascular resistance (PVR) was calculated as transpulmonary pressure gradient (mean PA pressure – PA wedge pressure), divided by CO and was expressed in Wood’s units (W.U.). During RHC, blood samples were obtained from the tip of free-floating catheter in pulmonary artery (PA) with deflated balloon (pulmonary arterial sample) and from wedged PA catheter (pulmonary venous sample), after discarding the initial 5 ml of aspirated blood. Wedged position was verified by fluoroscopy oximetry, whereby only samples with O_2_sat >90% were considered as representative of pulmonary venous blood^11^ (n=160). Cost considerations necessitated a more focused analysis for proteomic assays, so we identified patients with HFrEF and PVD as those subjects in the top quartile of PVR distribution (high PVR HF group, n=40), and we then contrasted them to subjects with HFrEF in the bottom quartile of PVR distribution (low PVR HF group, n=40). The patient disposition and flow are shown in the CONSORT diagram (Supplemental Figure 1).

### Targeted proteomic analysis

Blood samples from both sampling sites were handled in the same way and were placed into EDTA tubes and centrifuged within 20 min at a force of 800[×[g at a temperature of 4°C, distributed into aliquots, and frozen at −80°C until analysis. The Olink Target 96 Inflammation, Cardiovascular II and III panels (Olink Proteomics AB, Uppsala, Sweden) were used to measure proteins according to the manufacturer’s instructions. The Olink protocol utilizes the Proximity Extension Assay (PEA) technology, which allows for the simultaneous analysis of 92 analytes in very small sample (<10 μL) volumes, as described previously^12,16^. Briefly, pairs of oligonucleotide-labeled antibody probes specifically bind to their respective target proteins. When these probes come close to each other, the oligonucleotides hybridize, and proximity-dependent DNA polymerization occurs. This generates a unique polymerase chain reaction target sequence. The resulting DNA sequence is then detected and quantified using a microfluidic real-time polymerase chain reaction instrument (Biomark HD, Fluidigm, USA). To ensure data quality and account for variation between runs, internal controls including an extension control and an inter-plate control are used for normalization. The final output of the assay is presented as normalized protein expression (NPX) values, which are on a log2-scale. Higher normalized protein expression values indicate higher protein expression. Detailed assay validation data, such as detection limits and intra-assay and inter-assay precisions, can be found on the manufacturer’s website (www.olink.com). The list of proteins in all used OLINK panels is in the Supplemental table 1.

### Other biochemical analyses

Routine biochemistry was analyzed by an automated Abbott Architect ci1600 analyzer. Estimated glomerular filtration rate (eGFR) was calculated by the Chronic Kidney Disease Epidemiology Collaboration (CKD-EPI) equation. The B-type natriuretic peptide (BNP) concentrations were measured by using microparticle immunoassay (Architect BNP, Abbott Laboratories, Chicago, IL, USA; long-term analytical coefficient of variation [CV] 4.5%). Circulating IL-6 was verified by Elecsys IL-6 method (ECLIA by Roche Diagnostics, Mannheim, Germany; long-term analytical coefficient of variation [CV] 2.6%).

### Statistical analysis

Data were analyzed by using JMP18 software (SAS Institute, Inc., Cary, NC, USA). A two-sided p value of <0.05 was considered statistically significant. Data are presented as means ± SD or medians (interquartile ranges), where appropriate. Differences between groups were tested by using the chi-square test, unpaired Student t test, or analysis of variance. The statistical significance of pulmonary artery concentrations (to identify “markers”) or transpulmonary fold-change gradients (to identify putative “mediators”) was tested using unpaired and paired t test, respectively. The transpulmonary gradients were calculated as pulmonary postcapillary concentration (wedged PA catheter sampling) minus precapillary concentration (unwedged PA sampling). Differences in normalized protein expression (NPX; log2 fold change) were plotted against false-discovery rate (FDR)-adjusted p-values to obtain Volcano plots performed separately in control, high PVR HF and low PVR HF groups. P value < 0.05 was considered as significant. Protein-protein interaction network analysis was performed using STRING version 12.

## Results

Of the 160 patients with HF (median PVR: 2.88, IQR: 1.90-3.92 w.u., Supplemental Figure 2A) we performed transpulmonary proteomic analyses on those with low PVR (bottom quartile Q1, n=40, PVR<1.90 w.u.) and high PVR (top quartile, Q4, n=40, PVR>3.92 w.u.). Patients with HF were most often middle-aged males with typical characteristics of severely symptomatic HFrEF, while HF-free controls had similar anthropometric characteristics, but were slightly younger and more often female (Table 1).

**Table 1.**
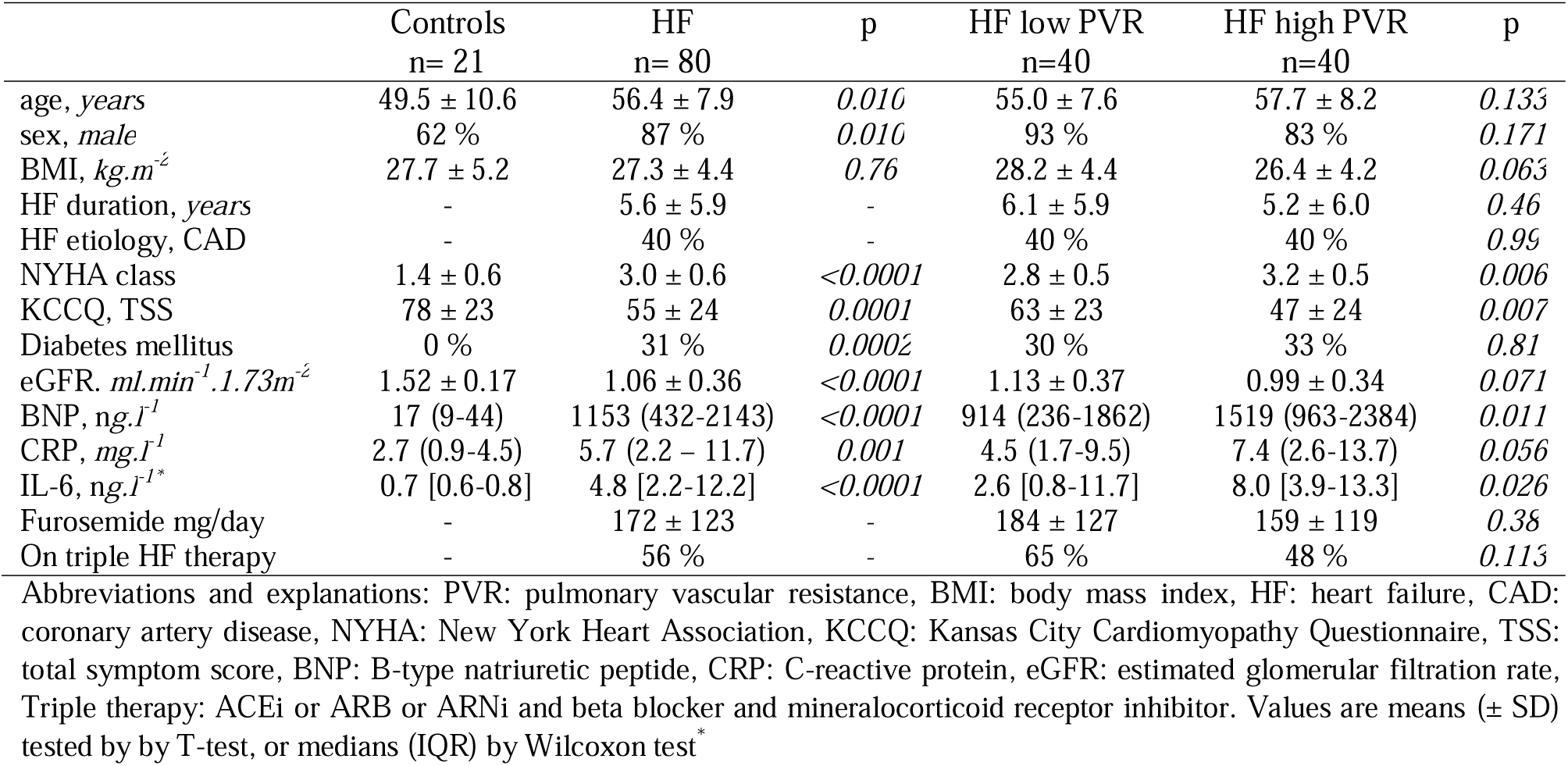
Baseline characteristics.

|  | Controls<br>n= 21 | HF<br>n= 80 | p | HF low PVR<br>n=40 | HF high PVR<br>n=40 | p |
| --- | --- | --- | --- | --- | --- | --- |
| age, years | 49.5 ± 10.6 | 56.4 ± 7.9 | 0.010 | 55.0 ± 7.6 | 57.7 ± 8.2 | 0.133 |
| sex, male | 62 % | 87 % | 0.010 | 93 % | 83 % | 0.171 |
| BMI, kg.m<sup>-2</sup> | 27.7 ± 5.2 | 27.3 ± 4.4 | 0.76 | 28.2 ± 4.4 | 26.4 ± 4.2 | 0.063 |
| HF duration, years | - | 5.6 ± 5.9 | - | 6.1 ± 5.9 | 5.2 ± 6.0 | 0.46 |
| HF etiology, CAD | - | 40 % | - | 40 % | 40 % | 0.99 |
| NYHA class | 1.4 ± 0.6 | 3.0 ± 0.6 | <0.0001 | 2.8 ± 0.5 | 3.2 ± 0.5 | 0.006 |
| KCCQ, TSS | 78 ± 23 | 55 ± 24 | 0.0001 | 63 ± 23 | 47 ± 24 | 0.007 |
| Diabetes mellitus | 0 % | 31 % | 0.0002 | 30 % | 33 % | 0.81 |
| eGFR, ml.min<sup>-1</sup>.1.73m<sup>-2</sup> | 1.52 ± 0.17 | 1.06 ± 0.36 | <0.0001 | 1.13 ± 0.37 | 0.99 ± 0.34 | 0.071 |
| BNP, ng.l<sup>-1</sup> | 17 (9-44) | 1153 (432-2143) | <0.0001 | 914 (236-1862) | 1519 (963-2384) | 0.011 |
| CRP, mg.l<sup>-1</sup> | 2.7 (0.9-4.5) | 5.7 (2.2 – 11.7) | 0.001 | 4.5 (1.7-9.5) | 7.4 (2.6-13.7) | 0.056 |
| IL-6, ng.l<sup>-1</sup>* | 0.7 [0.6-0.8] | 4.8 [2.2-12.2] | <0.0001 | 2.6 [0.8-11.7] | 8.0 [3.9-13.3] | 0.026 |
| Furosemide mg/day | - | 172 ± 123 | - | 184 ± 127 | 159 ± 119 | 0.38 |
| On triple HF therapy | - | 56 % | - | 65 % | 48 % | 0.113 |

Abbreviations and explanations: PVR: pulmonary vascular resistance, BMI: body mass index, HF: heart failure, CAD: coronary artery disease, NYHA: New York Heart Association, KCCQ: Kansas City Cardiomyopathy Questionnaire, TSS: total symptom score, BNP: B-type natriuretic peptide, CRP: C-reactive protein, eGFR: estimated glomerular filtration rate, Triple therapy: ACEi or ARB or ARNi and beta blocker and mineralocorticoid receptor inhibitor. Values are means (± SD) tested by T-test, or medians (IQR) by Wilcoxon test\*

Patients with HF had severely dilated and dysfunctional left ventricles, often with dysfunctional right ventricle, pulmonary hypertension (mean PA>20 mmHg in 86%) and reduced cardiac output (Table 2). Patients with HF and high or low PVR has similar anthropometry, age, sex, HF duration, HF etiology or HF therapy, but patients with PVD were more symptomatic (higher NYHA class, lower KCCQ scores), and had more severely elevated BNP, CRP and IL-6 levels compared with patients with HF and no PVD, along with more significant mitral regurgitation, higher PA pressures, and more severely reduced cardiac output. Importantly, high PVR patients had identical right atrial pressure, and had only marginally (mean difference 3.1 mmHg) higher PA wedge pressure, indicating that compensation and volume status were relatively well-matched.

**Table 2.**
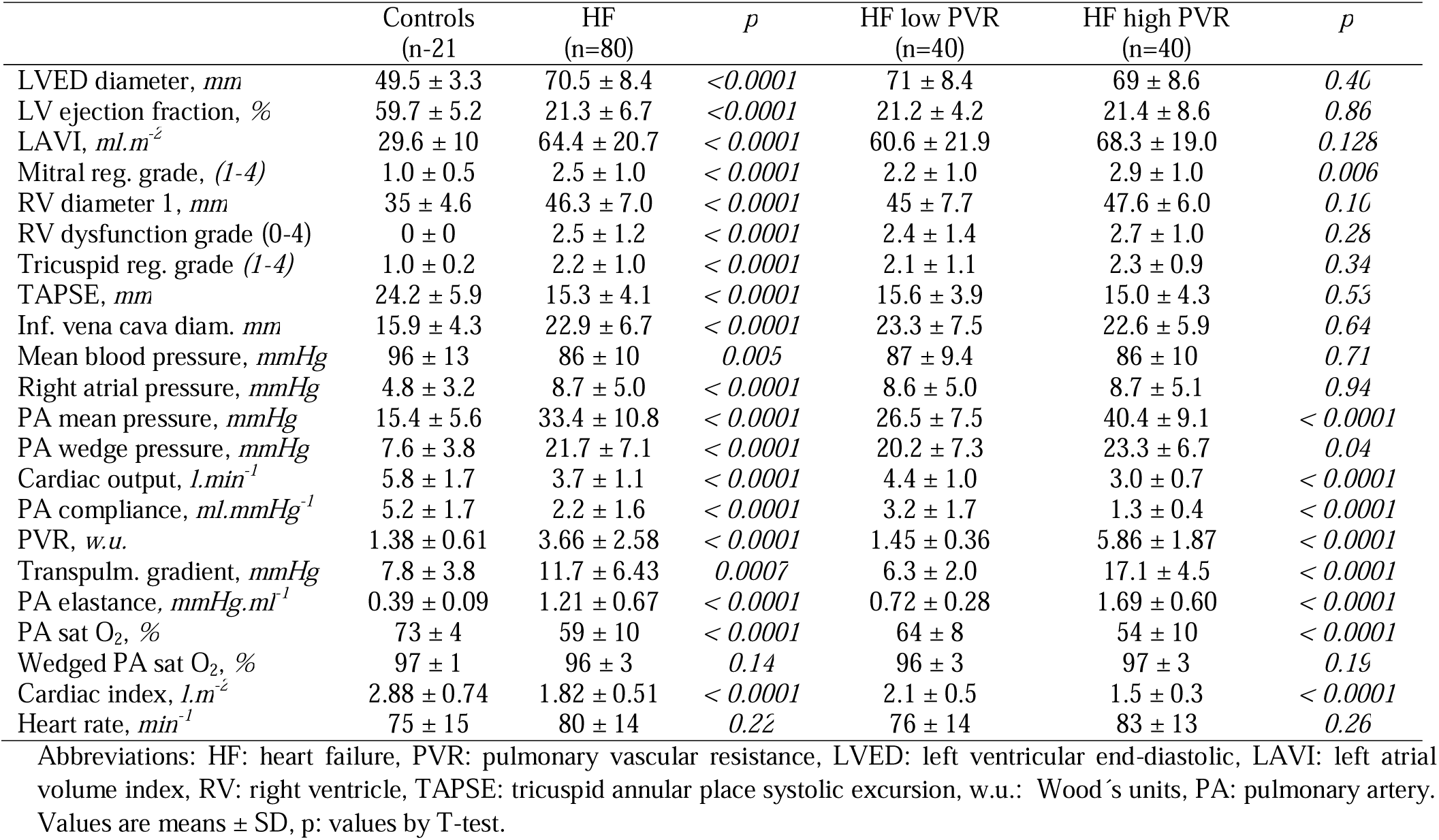
Echocardiography and invasive hemodynamics.

|  | Controls<br>(n=21) | HF<br>(n=80) | *p* | HF low PVR<br>(n=40) | HF high PVR<br>(n=40) | *p* |
| --- | --- | --- | --- | --- | --- | --- |
| LVED diameter, *mm* | 49.5 ± 3.3 | 70.5 ± 8.4 | <0.0001 | 71 ± 8.4 | 69 ± 8.6 | 0.40 |
| LV ejection fraction, % | 59.7 ± 5.2 | 21.3 ± 6.7 | <0.0001 | 21.2 ± 4.2 | 21.4 ± 8.6 | 0.86 |
| LAVI, *ml.m<sup>-2</sup>* | 29.6 ± 10 | 64.4 ± 20.7 | < 0.0001 | 60.6 ± 21.9 | 68.3 ± 19.0 | 0.128 |
| Mitral reg. grade, (1-4) | 1.0 ± 0.5 | 2.5 ± 1.0 | < 0.0001 | 2.2 ± 1.0 | 2.9 ± 1.0 | 0.006 |
| RV diameter 1, *mm* | 35 ± 4.6 | 46.3 ± 7.0 | < 0.0001 | 45 ± 7.7 | 47.6 ± 6.0 | 0.10 |
| RV dysfunction grade (0-4) | 0 ± 0 | 2.5 ± 1.2 | < 0.0001 | 2.4 ± 1.4 | 2.7 ± 1.0 | 0.28 |
| Tricuspid reg. grade (1-4) | 1.0 ± 0.2 | 2.2 ± 1.0 | < 0.0001 | 2.1 ± 1.1 | 2.3 ± 0.9 | 0.34 |
| TAPSE, *mm* | 24.2 ± 5.9 | 15.3 ± 4.1 | < 0.0001 | 15.6 ± 3.9 | 15.0 ± 4.3 | 0.53 |
| Inf. vena cava diam. *mm* | 15.9 ± 4.3 | 22.9 ± 6.7 | < 0.0001 | 23.3 ± 7.5 | 22.6 ± 5.9 | 0.64 |
| Mean blood pressure, *mmHg* | 96 ± 13 | 86 ± 10 | 0.005 | 87 ± 9.4 | 86 ± 10 | 0.71 |
| Right atrial pressure, *mmHg* | 4.8 ± 3.2 | 8.7 ± 5.0 | < 0.0001 | 8.6 ± 5.0 | 8.7 ± 5.1 | 0.94 |
| PA mean pressure, *mmHg* | 15.4 ± 5.6 | 33.4 ± 10.8 | < 0.0001 | 26.5 ± 7.5 | 40.4 ± 9.1 | < 0.0001 |
| PA wedge pressure, *mmHg* | 7.6 ± 3.8 | 21.7 ± 7.1 | < 0.0001 | 20.2 ± 7.3 | 23.3 ± 6.7 | 0.04 |
| Cardiac output, *l.min<sup>-1</sup>* | 5.8 ± 1.7 | 3.7 ± 1.1 | < 0.0001 | 4.4 ± 1.0 | 3.0 ± 0.7 | < 0.0001 |
| PA compliance, *ml.mmHg<sup>-1</sup>* | 5.2 ± 1.7 | 2.2 ± 1.6 | < 0.0001 | 3.2 ± 1.7 | 1.3 ± 0.4 | < 0.0001 |
| PVR, *w.u.* | 1.38 ± 0.61 | 3.66 ± 2.58 | < 0.0001 | 1.45 ± 0.36 | 5.86 ± 1.87 | < 0.0001 |
| Transpulm. gradient, *mmHg* | 7.8 ± 3.8 | 11.7 ± 6.43 | 0.0007 | 6.3 ± 2.0 | 17.1 ± 4.5 | < 0.0001 |
| PA elastance, *mmHg.ml<sup>-1</sup>* | 0.39 ± 0.09 | 1.21 ± 0.67 | < 0.0001 | 0.72 ± 0.28 | 1.69 ± 0.60 | < 0.0001 |
| PA sat O<sub>2</sub>, % | 73 ± 4 | 59 ± 10 | < 0.0001 | 64 ± 8 | 54 ± 10 | < 0.0001 |
| Wedge PA sat O<sub>2</sub>, % | 97 ± 1 | 96 ± 3 | 0.14 | 96 ± 3 | 97 ± 3 | 0.19 |
| Cardiac index, *l.m<sup>-2</sup>* | 2.88 ± 0.74 | 1.82 ± 0.51 | < 0.0001 | 2.1 ± 0.5 | 1.5 ± 0.3 | < 0.0001 |
| Heart rate, *min<sup>-1</sup>* | 75 ± 15 | 80 ± 14 | 0.22 | 76 ± 14 | 83 ± 13 | 0.26 |

Abbreviations: HF: heart failure, PVR: pulmonary vascular resistance, LVED: left ventricular end-diastolic, LAVI: left atrial volume index, RV: right ventricle, TAPSE: tricuspid annular place systolic excursion, w.u.: Wood's units, PA: pulmonary artery. Values are means ± SD, p: values by T-test.

### Proteomic biomarkers of heart failure or pulmonary vascular disease

First, we conducted targeted analysis of the proteome in unwedged PA blood, and compared all subjects with HF (high and low PVR together) to controls. A heat map of normalized protein expression (NPX) of HF vs controls showed clear separation of both groups (Supplemental Figure 3A). With FDR-adjusted p value <0.05 and cut-off of NPX log_2_- fold-change>2 or <0.5, we identified 33 proteins upregulated and 5 downregulated in HF (Suppl. Table 2 and Suppl Figure 3B). Upregulated proteins consisted of previously known markers of HF (NTproBNP, BNP, GDF15, FGF23, FGF21, for abbreviations, see Suppl. Table 1), inflammatory mediators (IL6, IL8, TNF receptor family members), chemokines (CCL15, CCL3, CCL20), and neurohormones (ACE2, IGFBP7, ADM, Renin). One of downregulated proteins in HF was the ROS defense protein paraoxonase (PON3).

Next, we compared the proteome of unwedged PA blood in patients with HF and high or low PVR to identify circulating biomarkers of PVD. We identified only 3 proteins that were significantly (FDR-adjusted p < 0.05) different in patients with HF and high PVR: Pulmonary Surfactant-associated Protein D (PSP-D), chemokine CCL3 and TNF receptor superfamily member TNFRSF13B.

### Transpulmonary proteomics: mediators of HF-related pulmonary vascular disease

As a next step, we analyzed transpulmonary protein gradients by subtracting concentrations in pulmonary venous blood (wedged PA sampling) from PA blood (unwedged PA sampling), assuming that proteins involved in pathogenesis of PVD are either taken up or elaborated across the pulmonary circulation. Results are presented in Table 3 and also as Volcano plots (Figure 1A-C). In controls, only one protein (the chemokine CCL19) displayed significant transpulmonary gradient (net uptake), which was of marginal significance (FDR-adjusted P value<0.05) (Figure 1A). In patients with HF and low PVR, we identified only 5 proteins with significant transpulmonary gradient – CCL19, Oncostatin M (OSM) and other two chemokines (Figure 1B).

**Figure 1:**
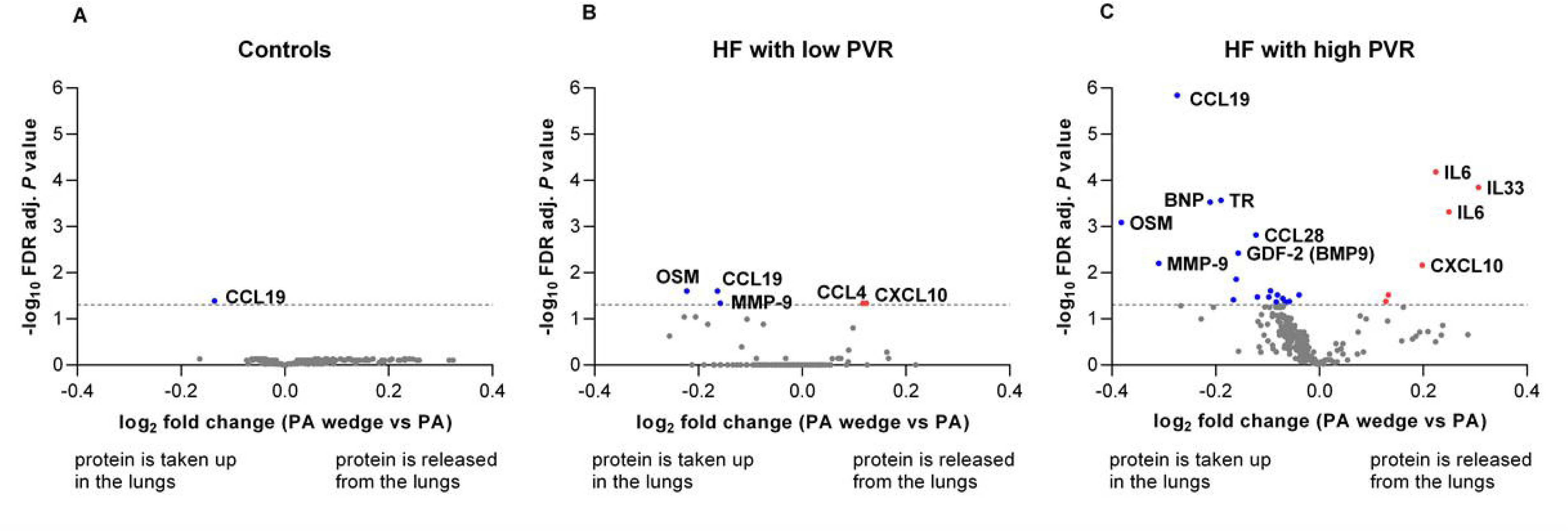
Volcano plots displaying proteins taken up in the lungs (blue dots) or released from the lungs (red dots) in Control subjects (A, n=21), in heart failure (HF) patients with low pulmonary vascular resistance (PVR) (B, n=40) and in HF with high PVR (C, n=40). Transpulmonary differences were calculated as postcapillary concentrations (pulmonary artery wedged blood) minus precapillary concentrations (pulmonary artery, PA).

**Table 3.** Significant transpulmonary gradients of proteins.

| Protein abbreviation | Protein name | UniProt ID | NPX K2-K1 (log2FC) | p-value Adjusted | Net pulmonary kinetics |
| --- | --- | --- | --- | --- | --- |
| **Controls** |  |  |  |  |  |
| CCL19 | C-C motif chemokine 19 | Q99731 | -0.13565 | 0.040787 | taken up |
| **Heart failure with low PVR** |  |  |  |  |  |
| OSM | Oncostatin M | P13725 | -0.22244 | 0.024964 | taken up |
| CCL19 | C-C motif chemokine 19 | Q99731 | -0.16323 | 0.024964 | taken up |
| MMP9 | Matrix metalloproteinase-9 | P14780 | -0.15822 | 0.046083 | taken up |
| CCL4 | C-C motif chemokine 4 | P13236 | 0.116997 | 0.046083 | released |
| CXCL10 | C-X-C motif chemokine 10 |  | 0.123701 | 0.046083 | released |
| **Heart failure with high PVR** |  |  |  |  |  |
| OSM | Oncostatin M | P13725 | -0.38186 | 0.000815 | taken up |
| MMP9 | Matrix metalloproteinase 9 | P14780 | -0.31003 | 0.006239 | taken up |
| CCL19 | C-C motif chemokine 19 | Q99731 | -0.27428 | 0.000001 | taken up |
| BNP | B-type natriuretic peptide | P16860 | -0.21109 | 0.000298 | taken up |
| TR (TFCR) | Transferrin receptor protein 1 | P02786 | -0.19005 | 0.000271 | taken up |
| IL2RB | IL-2 receptor subunit beta | P14784 | -0.16617 | 0.038841 | taken up |
| TGFA | Transforming growth factor alpha | P01135 | -0.16067 | 0.013826 | taken up |
| GDF2 (BMP9) | Growth differentiation factor 2 (Bone morphogenetic protein 9) | Q9UK05 | -0.15676 | 0.003769 | taken up |
| CCL28 | C-C motif chemokine 28 | Q9NRJ3 | -0.12256 | 0.001523 | taken up |
| GT (FABP6) | Gastrotropin (Fatty Acid Binding Protein 6) | P51161 | -0.11972 | 0.033794 | taken up |
| MERTK | Tyrosine-protein kinase Mer | Q12866 | -0.09758 | 0.033794 | taken up |
| FABP2 | Fatty acid-binding protein, intestinal | P12104 | -0.09427 | 0.024708 | taken up |
| CCL25 | C-C motif chemokine 25 | O15444 | -0.08313 | 0.042691 | taken up |
| KIM1 (HAVCR1) | Kidney injury molecule 1 (Hepatitis A virus cellular receptor 1) | Q96D42 | -0.08123 | 0.030517 | taken up |
| TF (TFRC) | Transferrin receptor protein 1 | P02786 | -0.07098 | 0.036286 | taken up |
| SCF (KITLG)* | Stem Cell Factor (Kit Ligand) | P21583 | -0.06656 | 0.042691 | taken up |
| TIE2 (TEK) | Angiopoietin-1 receptor | Q02763 | -0.05807 | 0.041734 | taken up |
| PRELP | Prolargin | P51888 | -0.0393 | 0.030517 | taken up |
| CCL4 | C-C motif chemokine 4 | P13236 | 0.127964 | 0.041734 | released |
| MCP-3 (CCL7) | Monocyte chemotactic protein 3 (C-C motif chemokine 7) | P80098 | 0.132624 | 0.030517 | released |
| CXCL10 | C-X-C motif chemokine 10 | P02778 | 0.198088 | 0.006899 | released |
| IL6 * | Interleukin 6 | P05231 | 0.224349 | 6.59E-05 | released |
| IL6 ** | Interleukin 6 | P05231 | 0.249581 | 0.000483 | released |
| IL33 | Interleukin 33 | O95760 | 0.306348 | 0.000142 | released |

In brackets are newer abbreviations (from web https://olink.com/products/olink-target-96, in suppl 1).
For Uniprot ID see supplement 1. \* in Olink panel CVII, \*\* in Olink panel Inflammation.

In contrast, patients with HF and high PVR displayed 23 proteins with significant transpulmonary gradients, including 18 proteins that were consumed and 5 proteins being released from the lung into circulation (Figure 1C), mostly inflammatory cytokines and chemokines. The highest lung uptake was observed for chemokine CCL19, cytokine Oncostatin M, BNP, Bone morphogenic protein 9 (BMP9 aka GDF2), Transforming growth factor alfa (TGFA), Transferrin receptor (TR) and matrix metalloproteinase 9 (MMP9) (Figure 2A,B). The highest lung release was observed for interleukin 6 and 33 (IL6, IL33) and the chemokines MCP3, CCL4 and CXCL10. Protein ontology analysis using STRING demonstrated central roles of IL6, OSM and CCL4 in patients with HF and high PVR (Suppl. Figure 4).

**Figure 2:**
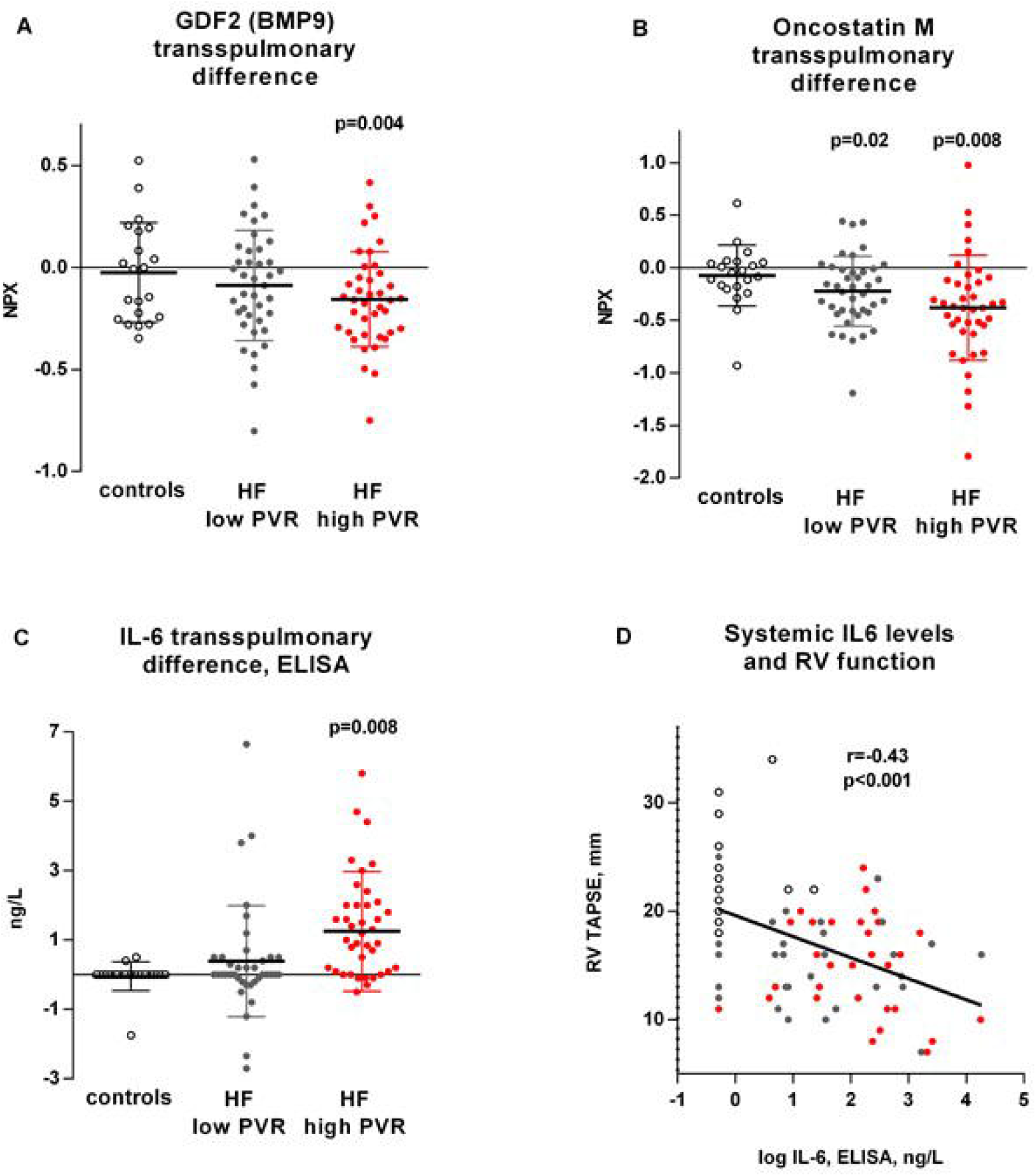
Individual transpulmonary differences in Control subjects (open dots, n=21), in heart failure (HF) patients with low pulmonary vascular resistance (PVR) (grey dots, n=40) and in HF with high PVR (red dots, n=40) of protein Growth differentiation factor 2 (GDF2, aka BMP9, A), Oncostatin M (B), and Interleukin 6 (IL-6). Values in A, B are from PEA Olink assay and expressed as normalized protein expression units (NPX), values in C are by ELISA and expressed as ng/L. P values (A-C) are false discovery rate (FDR)-adjusted and represent significant difference from net zero gradient (null hypothesis). Relation of RV function (tricuspid anulus plane systolic excursion, TAPSE) with mixed venous concentration of IL-6, correlation coefficient and linear regression line (D).

Distribution analyses in the most relevant hits excluded transpulmonary differences being driven by outlying values (Figure 2). Because several proteins (IL6, CCL3) are present in both OLINK panels (Inflammation and CVII), we were able to assess reproducibility of the method. As shown in Suppl. Figure 1D, results of both IL6 measurements by OLINK correlated very well (r=0.99) and both IL6 dots in Volcano plots in Figure 1C were close to each other. We further validated BNP results by ELISA measurements and observed excellent correlation for BNP (r=0.97, p<0.0001), and acceptable for IL6 (r=0.67, p<0.001). There was also significantly increased IL6 release from lungs of HF patients with high PVR based upon ELISA measurements (Figure 2C), and systemic IL6 concentration by ELISA correlated with right ventricular tricuspid annulus plane systolic excursion (RV TAPSE), indicating the association of IL6 levels with poorer RV function (Figure 2D).

## Discussion

This is the first study to systematically characterize the transpulmonary proteome in HF-related pulmonary vascular disease (PVD). The principal findings are that transpulmonary proteomic signatures of patients with PVD strongly differ from controls and from patients with HF and lower PVR, particularly for gradients of inflammatory chemokines, cytokines from IL6 superfamily, and by proteins that are involved in pulmonary vascular remodeling typical for G1 PH. This study supports the observations that in considerable number of patients, there is a phenotype overlap between G1 and G2 PH^17,18^. The present data raise a new hypothesis that PVD in HF is caused by vascular inflammation and structural vessel remodeling, in addition to vasoconstriction, which may be important therapeutic targets to treat or prevent PVD in HF.

Pulmonary vascular disease in HF adds a hemodynamic load in excess of that transmitted passively across the lungs from left atrial hypertension. Increases in PVR in HF are independently associated with increased mortality^1,19^, and impose an additional load to RV and promote progression of HF,^3^ while also contributing to gas exchange abnormalities and pathologic increases in lung congestion^20^. Increase in PVR in G2 PH develops in part due to vasoconstriction, as it is at least partly reversible with pulmonary vasodilators in most patients^17^, by the direct effect of congestion on pulmonary vessels^2,21^ and through chronic structural alterations of pulmonary vasculature, such as vessel thickening and rarefaction^3,4,6^.

Using a biomarker approach comparing mixed venous blood samples of patients with high and low PVR, we identified Surfactant Protein D (PSP-D) as a biomarker associated with high PVR in HF. Elevated PSP-D in HF likely reflects stressed or disrupted alveolar-capillary interface,^3^ as plasma levels of PSD-D are known to be elevated for weeks after resolution of cardiogenic pulmonary edema^22,23^. Because transpulmonary gradients were net zero, elevated PSD-D is a likely a biomarker of previous episodes of congestion, rather than actively contributing to PVD pathophysiology, at least at steady state and in the compensated state as evaluated herein.

Analyses of transpulmonary proteomic gradients provided a number of novel findings. The results indicate that pulmonary vascular homeostasis in heathy subjects is characterized by almost zero transpulmonary differences of regulatory proteins. In patients with HF, with increasing PVR we observed transpulmonary gradients of several proinflammatory cytokines and chemokines and also proteins linked to vascular remodeling in PAH. Inflammation have been associated with G1 PH^24^, but we show here that it is also associated with G2 PH, alongside with other non-inflammatory mechanisms shared with G1 PH. Importantly, we studied HF subjects after intravenous decongestion - so patients with and without PVD had identical central venous pressure so the effect of congestion in our study was effectively mitigated and observed transpulmonary gradients are representative of chronic processes of PVD.

### Inflammatory mediators associated with pulmonary vascular disease

Specifically, elevated PVR in HF was associated with net lung release of interleukin 6 (IL6). Patients with HF, and particularly those with PVD, had greater systemic inflammation from venous blood samples, as reflected by IL6 and CRP. Our transpulmonary data suggest that the lungs themselves are a source of inflammatory mediators and IL6 release in HF with PVD. Our findings also strongly support causal link between IL6 and pulmonary hypertension, that was already suggested in patients with PAH^25^ and in a PH-HFpEF experimental model^26^. The inflammatory mediators may originate from pulmonary vessels, from perivascular inflammatory infiltrates, as observed in PAH^27^, or from extravascular cells such as alveolar myofibroblasts responding to capillary stress failure,^28^ or areas of lung interstitial inflammation corresponding to ground glass opacities detected by computer tomography in some G2 or G3 PH patients^17^.

Positive IL6 transpulmonary gradient in HF patients comprised ∼8% of systemic venous blood IL6 levels, indicating that pulmonary IL6 production contributes considerably to the circulating pool of IL6. This may be relevant for systemic inflammatory response in HF, but this does not necessarily mean that IL6 itself is causing the increase of PVR. In the TRANSFORM-UK trial in G1 PH, the IL6 receptor antagonist tocilizumab reduced CRP levels, but did not reduce PVR, and actually increased IL6 levels^29^. Lung IL6 release may be a response to upstream vascular injury, rather than being a causal factor in human PH,^29^ but it also possible that IL6 contributes to pulmonary vascular disease through remodeling or other effects. The HERMES and ATHENA trials are currently testing the effects of the IL-6 targeting antibody ziltivekimab on reducing worsening HF or cardiovascular death, as well as functional outcomes in patients with HFmrEF and HFpEF and systemic inflammation (NCT05636176 and NCT06200207). It may be that ziltivekimab works in part through reduction of pulmonary vascular inflammation or has effects on pulmonary vascular loading or right heart function.

Another inflammatory mediator secreted from lungs with high PVR is interleukin 33, an alarmin previously implicated in hyperproliferative response of pulmonary vessels in hypoxic PH^30^. Interestingly, the receptor for IL33 is ST2 protein, and circulating soluble ST2 protein levels are associated with proinflammatory comorbidities, right ventricular pressure overload and dysfunction in HF^23^, probably reflecting increased IL33 lung release^31^. The lungs of patients with HF and high PVR also secreted chemokine CXCL10, which impairs pulmonary angiogenesis,^32^ and is elevated in HF^33^, as well as chemokine CCL4 (formerly MIP 1β), also known to be upregulated in the lungs of individuals with PH^32^.

A number of proteins were highly consumed in the high PVR lungs (i.e. with negative gradient), including Oncostatin M (OSM). OSM is pleotropic cytokine belonging to IL6 family that signals via GP130 coreceptor and regulates inflammation and endothelial IL6 production. In an experimental model, lung OSM administration induces recruitment of inflammatory cells into the lungs, leading to a profibrotic gene expression footprint, collagen deposition, and pathological correlates characteristic of human interstitial lung disease or lung changes due to systemic scleroderma^34^. It was shown that OSM induces IL6 secretion from endothelial cells^35^ and smooth muscle cells^36^, and also stimulates pulmonary release of IL33^37,38^. Therefore, the increased transpulmonary uptake of OSM observed here in high PVR lungs could be a “primary upstream event”, leading to pulmonary inflammation and release of other secondary cytokines, such as IL6 and IL33. OSM could therefore also serve as a target to treat PVD. A monoclonal antibody against OSM vixarelimab is currently being tested for pulmonary fibrotic disease (NCT05785624), and the present data suggest that effects on PVD should be examined as well.

Increased CCL19 and MMP9 uptake in the high PVR group may reflect leucocyte recruitment, inflammation-triggered ECM remodeling with degradation of elastin, endothelial to mesenchymal transition (EMT), and thickening of pulmonary vascular walls^39,40^. Similarly, chemokine CCL28 displayed increased uptake in the high PVR group, and was previously identified as a marker of PH in patients with chronic obstructive pulmonary disease^41^.

### Non-inflammatory mediators associated with PVD

Interestingly, we observed enhanced lung uptake of Growth differentiation factor 2 (GDF2, also known as BMP9) in high PVR group. GDF2 is produced in the liver, and is one of key ligands of bone morphogenic protein receptor (BMPR2), mutations of which are the most common cause of hereditary G1 PH^5,42^. Acquired deficiency of GDF2 due to liver dysfunction is causally linked to the development of portopulmonary hypertension^43^ and hepatopulmonary syndrome^44^. We cannot make inferences as to whether enhanced uptake of GDF2 in the high PVR group was causally related to high PVR, or it represent a compensatory response to antagonize profibrotic TGF/activin signaling^5^. However, the present results suggest that TGF/activin/BMP signaling pathway, which drives structural remodeling of pulmonary vessels in G1 PH, is also part of G2 PH pathophysiology, and could be a drug target. The activin receptor ligand trap sotatercept successfully prevented pulmonary remodeling in experimental model of PVD due to left heart disease^45^ and is currently clinically tested in PH-HFpEF patients with significant precapillary component in the CADENCE trial (NCT04945460).

Finally, we observed enhanced uptake of B-type natriuretic peptide (BNP) across the lungs in patients with HF and high PVR. This likely correspond to enhanced contra-regulatory vasodilatation via cGMP pathway in high PVR HF subjects^10^ which may explain why inhibition of BNP degradation by neprilysin inhibition improves PVR in HFrEF^46^.

This study must be interpreted in the context of several limitations. The study was prospective but cross-sectional, limiting the ability to reach conclusion regarding causality. Due to budget constraints, we compared two outlying quartiles rather than all patients with HF, yet both HF groups were balanced in term of relative congestion, comorbidities, age, and anthropometrics. Notably, PEA technology uses similar optimized double antibody detection like conventional sandwich ELISA, but offers larger sensitivity and range - so the position of ELISA as a gold standard is questionable, though we did confirm findings for IL6 and BNP by ELISA. PEA technology does not report absolute protein concentration, but this was not an obstacle for the design of our study that looked at relative changes of the analytes across the lungs, providing internal normalization.

In summary, the lungs of patients with HF, particularly those with high PVR, display abnormal uptake and release of inflammatory chemokines and cytokines from IL6 family, along with increased pulmonary uptake of Oncostatin M and GDF2, a mediator of lung inflammation and a ligand of the BMPR2 receptor involved in pathogenesis of G1 PH, respectively. These data provide novel insights into the mechanisms underlying development of PVD in HF, and call for further evaluation in clinical trials targeting these pathways to improve outcomes in patients with PVD due to HF.

## Acknowledgements

Special thanks to Tomas and Veronika Veberovi, Cizkrajice.

## Data availability statement

The data are available upon reasonable request.

## References

1. Maron BA, Brittain EL, Hess E, Waldo SW, Baron AE, Huang S, et al. Pulmonary vascular resistance and clinical outcomes in patients with pulmonary hypertension: a retrospective cohort study. Lancet Respir Med. 2020;8:873–884. doi: 10.1016/S2213-2600(20)30317-9

2. Melenovsky V, Andersen MJ, Andress K, Reddy YN, Borlaug BA. Lung congestion in chronic heart failure: haemodynamic, clinical, and prognostic implications. Eur J Heart Fail. 2015;17:1161–1171. doi: 10.1002/ejhf.417

3. Huston JH, Shah SJ. Understanding the Pathobiology of Pulmonary Hypertension Due to Left Heart Disease. Circ Res. 2022;130:1382–1403. doi: 10.1161/CIRCRESAHA.122.319967

4. Fayyaz AU, Edwards WD, Maleszewski JJ, Konik EA, DuBrock HM, Borlaug BA, et al. Global Pulmonary Vascular Remodeling in Pulmonary Hypertension Associated With Heart Failure and Preserved or Reduced Ejection Fraction. Circulation. 2018;137:1796–1810. doi: 10.1161/CIRCULATIONAHA.117.031608

5. Guignabert C, Aman J, Bonnet S, Dorfmuller P, Olschewski AJ, Pullamsetti S, et al. Pathology and pathobiology of pulmonary hypertension: current insights and future directions. Eur Respir J. 2024;64. doi: 10.1183/13993003.01095-2024

6. Fayyaz AU, Sabbah MS, Dasari S, Griffiths LG, DuBrock HM, Wang Y, et al. Histologic and proteomic remodeling of the pulmonary veins and arteries in a porcine model of chronic pulmonary venous hypertension. Cardiovasc Res. 2023;119:268-282. doi: 10.1093/cvr/cvac005

7. Maron BA, Bortman G, De Marco T, Huston JH, Lang IM, Rosenkranz SH, et al. Pulmonary hypertension associated with left heart disease. Eur Respir J. 2024;64. doi: 10.1183/13993003.01344-2024

8. Murashige D, Jang C, Neinast M, Edwards JJ, Cowan A, Hyman MC, et al. Comprehensive quantification of fuel use by the failing and nonfailing human heart. Science. 2020;370:364-368. doi: 10.1126/science.abc8861

9. Monzo L, Sedlacek K, Hromanikova K, Tomanova L, Borlaug BA, Jabor A, et al.. Myocardial ketone body utilization in patients with heart failure: The impact of oral ketone ester. Metabolism. 2021;115:154452. doi: 10.1016/j.metabol.2020.154452

10. Melenovsky V, Al-Hiti H, Kazdova L, Jabor A, Syrovatka P, Malek I, et al. Transpulmonary B-type natriuretic peptide uptake and cyclic guanosine monophosphate release in heart failure and pulmonary hypertension: the effects of sildenafil. J Am Coll Cardiol. 2009;54:595–600. doi: 10.1016/j.jacc.2009.05.021

11. Tang WHW, Wilcox JD, Jacob MS, Rosenzweig EB, Borlaug BA, Frantz RP, et al. Comprehensive Diagnostic Evaluation of Cardiovascular Physiology in Patients With Pulmonary Vascular Disease: Insights From the PVDOMICS Program. Circ Heart Fail. 2020;13:e006363. doi: 10.1161/CIRCHEARTFAILURE.119.006363

12. Wik L, Nordberg N, Broberg J, Bjorkesten J, Assarsson E, Henriksson S, et al. Proximity Extension Assay in Combination with Next-Generation Sequencing for High-throughput Proteome-wide Analysis. Mol Cell Proteomics. 2021;20:100168. doi: 10.1016/j.mcpro.2021.100168

13. Said SI. Metabolic functions of the pulmonary circulation. Circ Res. 1982;50:325–333. doi: 10.1161/01.res.50.3.325

14. Eldjarn GH, Ferkingstad E, Lund SH, Helgason H, Magnusson OT, Gunnarsdottir K, et al. Large-scale plasma proteomics comparisons through genetics and disease associations. Nature. 2023;622:348–358. doi: 10.1038/s41586-023-06563-x

15. Rosenkranz S, Preston IR. Right heart catheterisation: best practice and pitfalls in pulmonary hypertension. Eur Respir Rev. 2015;24:642–652. doi: 10.1183/16000617.0062-2015

16. Assarsson E, Lundberg M, Holmquist G, Bjorkesten J, Thorsen SB, Ekman D, et al. Homogenous 96-plex PEA immunoassay exhibiting high sensitivity, specificity, and excellent scalability. PLoS One. 2014;9:e95192. doi: 10.1371/journal.pone.0095192

17. Borlaug BA, Larive B, Frantz RP, Hassoun P, Hemnes A, Horn E, et al. Pulmonary hypertension across the spectrum of left heart and lung disease. Eur J Heart Fail. 2024;26:1642–1651. doi: 10.1002/ejhf.3302

18. Hemnes AR, Leopold JA, Radeva MK, Beck GJ, Abidov A, Aldred MA, et al. Clinical Characteristics and Transplant-Free Survival Across the Spectrum of Pulmonary Vascular Disease. J Am Coll Cardiol. 2022;80:697–718. doi: 10.1016/j.jacc.2022.05.038

19. Miller WL, Grill DE, Borlaug BA. Clinical features, hemodynamics, and outcomes of pulmonary hypertension due to chronic heart failure with reduced ejection fraction: pulmonary hypertension and heart failure. JACC Heart Fail. 2013;1:290–299. doi: 10.1016/j.jchf.2013.05.001

20. Omote K, Sorimachi H, Obokata M, Reddy YNV, Verbrugge FH, Omar M,et al.. Pulmonary vascular disease in pulmonary hypertension due to left heart disease: pathophysiologic implications. Eur Heart J. 2022;43:3417–3431. doi: 10.1093/eurheartj/ehac184

21. West JB, Dollery CT, Heard BE. Increased Pulmonary Vascular Resistance in the Dependent Zone of the Isolated Dog Lung Caused by Perivascular Edema. Circ Res. 1965;17:191–206. doi: 10.1161/01.res.17.3.191

22. De Pasquale CG, Arnolda LF, Doyle IR, Grant RL, Aylward PE, Bersten AD. Prolonged alveolocapillary barrier damage after acute cardiogenic pulmonary edema. Crit Care Med. 2003;31:1060–1067. doi: 10.1097/01.CCM.0000059649.31659.22

23. Guazzi M, Novello G, Bursi F, Caretti A, Furlotti N, Arena R, et al.. Biomarkers of lung congestion and injury in acute heart failure. ESC Heart Fail. 2024. doi: 10.1002/ehf2.14982

24. Rabinovitch M, Guignabert C, Humbert M, Nicolls MR. Inflammation and immunity in the pathogenesis of pulmonary arterial hypertension. Circ Res. 2014;115:165–175. doi: 10.1161/CIRCRESAHA.113.301141

25. Simpson CE, Chen JY, Damico RL, Hassoun PM, Martin LJ, Yang J, et al. Cellular sources of interleukin-6 and associations with clinical phenotypes and outcomes in pulmonary arterial hypertension. Eur Respir J. 2020;55. doi: 10.1183/13993003.01761-2019

26. Ranchoux B, Nadeau V, Bourgeois A, Provencher S, Tremblay E, Omura J, et al. Metabolic Syndrome Exacerbates Pulmonary Hypertension due to Left Heart Disease. Circ Res. 2019;125:449–466. doi: 10.1161/CIRCRESAHA.118.314555

27. Liu SF, Nambiar Veetil N, Li Q, Kucherenko MM, Knosalla C, Kuebler WM. Pulmonary hypertension: Linking inflammation and pulmonary arterial stiffening. Front Immunol. 2022;13:959209. doi: 10.3389/fimmu.2022.959209

28. Jasmin JF, Calderone A, Leung TK, Villeneuve L, Dupuis J. Lung structural remodeling and pulmonary hypertension after myocardial infarction: complete reversal with irbesartan. Cardiovasc Res. 2003;58:621–631. doi: 10.1016/s0008-6363(03)00290-6

29. Toshner M, Church C, Harbaum L, Rhodes C, Villar Moreschi SS, Liley J, et al. Mendelian randomisation and experimental medicine approaches to interleukin-6 as a drug target in pulmonary arterial hypertension. Eur Respir J. 2022;59. doi: 10.1183/13993003.02463-2020

30. Indralingam CS, Gutierrez-Gonzalez AK, Johns SC, Tsui T, Cannon DT, Fuster MM,et al.. IL-33/ST2 receptor-dependent signaling in the development of pulmonary hypertension in Sugen/hypoxia mice. Physiol Rep. 2022;10:e15185. doi: 10.14814/phy2.15185

31. AbouEzzeddine OF, McKie PM, Dunlay SM, Stevens SR, Felker GM, Borlaug BA, et al. Suppression of Tumorigenicity 2 in Heart Failure With Preserved Ejection Fraction. J Am Heart Assoc. 2017;6. doi: 10.1161/JAHA.116.004382

32. Mamazhakypov A, Viswanathan G, Lawrie A, Schermuly RT, Rajagopal S. The role of chemokines and chemokine receptors in pulmonary arterial hypertension. Br J Pharmacol. 2021;178:72–89. doi: 10.1111/bph.14826

33. Altara R, Manca M, Hessel MH, Gu Y, van Vark LC, Akkerhuis KM, et al.. CXCL10 Is a Circulating Inflammatory Marker in Patients with Advanced Heart Failure: a Pilot Study. J Cardiovasc Transl Res. 2016;9:302–314. doi: 10.1007/s12265-016-9703-3

34. Mozaffarian A, Brewer AW, Trueblood ES, Luzina IG, Todd NW, Atamas SP, et al. Mechanisms of oncostatin M-induced pulmonary inflammation and fibrosis. J Immunol. 2008;181:7243–7253. doi: 10.4049/jimmunol.181.10.7243

35. Brown TJ, Rowe JM, Liu JW, Shoyab M. Regulation of IL-6 expression by oncostatin M. J Immunol. 1991;147:2175–2180.

36. Bernard C, Merval R, Lebret M, Delerive P, Dusanter-Fourt I, Lehoux S, et al. Oncostatin M induces interleukin-6 and cyclooxygenase-2 expression in human vascular smooth muscle cells : synergy with interleukin-1beta. Circ Res. 1999;85: 1124–1131. doi: 10.1161/01.res.85.12.1124

37. Richards CD, Izakelian L, Dubey A, Zhang G, Wong S, Kwofie K, et al. Regulation of IL-33 by Oncostatin M in Mouse Lung Epithelial Cells. Mediators Inflamm. 2016;2016:9858374. doi: 10.1155/2016/9858374

38. Botelho F, Dubey A, Ayaub EA, Park R, Yip A, Humbles A,et al. IL-33 Mediates Lung Inflammation by the IL-6-Type Cytokine Oncostatin M. Mediators Inflamm. 2020;2020:4087315. doi: 10.1155/2020/4087315

39. Thenappan T, Chan SY, Weir EK. Role of extracellular matrix in the pathogenesis of pulmonary arterial hypertension. Am J Physiol Heart Circ Physiol. 2018;315:H1322–H1331. doi: 10.1152/ajpheart.00136.2018

40. Stenmark KR, Frid M, Perros F. Endothelial-to-Mesenchymal Transition: An Evolving Paradigm and a Promising Therapeutic Target in PAH. Circulation. 2016;133:1734–1737. doi: 10.1161/CIRCULATIONAHA.116.022479

41. Zhang Y, Lin P, Hong C, Jiang Q, Xing Y, Tang X, et al. Serum cytokine profiles in patients with chronic obstructive pulmonary disease associated pulmonary hypertension identified using protein array. Cytokine. 2018;111:342–349. doi: 10.1016/j.cyto.2018.09.005

42. Long L, Ormiston ML, Yang X, Southwood M, Graf S, Machado RD, et al. Selective enhancement of endothelial BMPR-II with BMP9 reverses pulmonary arterial hypertension. Nat Med. 2015;21:777–785. doi: 10.1038/nm.3877

43. Nikolic I, Yung LM, Yang P, Malhotra R, Paskin-Flerlage SD, Dinter T, et al. Bone Morphogenetic Protein 9 Is a Mechanistic Biomarker of Portopulmonary Hypertension. Am J Respir Crit Care Med. 2019;199:891–902. doi: 10.1164/rccm.201807-1236OC

44. Robert F, Certain MC, Baron A, Thuillet R, Duhaut L, Ottaviani M, et al. Disrupted BMP-9 Signaling Impairs Pulmonary Vascular Integrity in Hepatopulmonary Syndrome. Am J Respir Crit Care Med. 2024;210:648–661. doi: 10.1164/rccm.202307-1289OC

45. Joshi SR, Atabay EK, Liu J, Ding Y, Briscoe SD, Alexander MJ, et al.. Sotatercept analog improves cardiopulmonary remodeling and pulmonary hypertension in experimental left heart failure. Front Cardiovasc Med. 2023;10:1064290. doi: 10.3389/fcvm.2023.1064290

46. Zern EK, Cheng S, Wolfson AM, Hamilton MA, Zile MR, Solomon SD, et al. Angiotensin Receptor-Neprilysin Inhibitor Therapy Reverses Pulmonary Hypertension in End-Stage Heart Failure Patients Awaiting Transplantation. Circ Heart Fail. 2020;13:e006696. doi: 10.1161/CIRCHEARTFAILURE.119.006696

