## Supplementary material for "Transpulmonary proteome gradients identify pathways involved in the development of pulmonary vascular disease in heart failure": Suppl Table 2

**Supplemental table 2 Biomarkers in PA blood**

| Protein  abbreviation | Protein name | UniProt  ID | log2fold change (FC) | p-value  Adjusted |
| --- | --- | --- | --- | --- |
| **Biomarkers of HF** (Controls vs HF comparison) | | | | |
| BNP*** | Brain natriuretic peptide (N-terminal prohormone of BNP) | P16860 | 5.105416 | 1.58E-20 |
| BNP* | Brain natriuretic peptide | P16860 | 5.623247 | 7.45E-17 |
| GDF15 | Growth differentiation factor 15 | Q99988 | 2.088447 | 2.39E-16 |
| FGF23* | Fibroblast growth factor 23 | Q9GZV9 | 2.673613 | 7.68E-16 |
| FGF23** | Fibroblast growth factor 23 | Q9GZV9 | 3.181233 | 3.37E-15 |
| IL8 (CXCL8) | Interleukin 8 (CXCL8) | P10145 | 1.593858 | 5.71E-14 |
| ACE2 | Angiotensin-converting enzyme 2 | Q9BYF1 | 1.468633 | 6.43E-13 |
| IGFBP7 | Insulin-like growth factor-binding protein 7 | Q16270 | 1.16157 | 1.51E-12 |
| TNFRSF10B (TRAIL-R2) | TNF receptor superfamily member 10B (TNF-related apoptosis-inducing ligand receptor 2) | O14763 | 1.229691 | 7.55E-12 |
| TFF3 | Trefoil factor 3 | Q07654 | 1.103298 | 9.67E-12 |
| TNFRSF11A | TNF receptor superfamily member 11A | Q9Y6Q6 | 1.01974 | 2.38E-11 |
| REN | Renin | P00797 | 1.98645 | 2.79E-11 |
| JAM-A (F11R) | Junctional adhesion molecule A | Q9Y624 | 1.061488 | 3.26E-11 |
| IL6* | Interleukin 6 | P05231 | 1.615907 | 6.29E-11 |
| IL6** | Interleukin 6 | P05231 | 1.839921 | 7.52E-11 |
| CCL15 | C-C motif chemokine 15 | Q16663 | 1.221489 | 8.47E-11 |
| FABP4 | Fatty acid-binding protein, adipocyte | P15090 | 2.317406 | 4.48E-10 |
| ADM | Pro-adrenomedullin | P35318 | 1.0082 | 7.15E-10 |
| CCL3** | C-C motif chemokine 3 (Macrophage inflammatory protein 1-alpha) | P10147 | 1.236451 | 3.11E-09 |
| MMP12 | Macrophage metalloelastase | P39900 | 1.089928 | 4.52E-09 |
| VSIG2 | V-set and immunoglobulin domain-containing protein 2 | Q96IQ7 | 1.021618 | 1.85E-08 |
| CSTB | Cystatin-B | P04080 | 1.02126 | 1.4E-07 |
| HAVCR1 (KIM1) | Hepatitis A virus cellular receptor 1 (Kidney Injury Molecule -1) | Q96D42 | 1.292431 | 1.67E-07 |
| CCL3* | C-C motif chemokine 3 (Macrophage inflammatory protein 1-alpha) | P10147 | 1.118741 | 4.46E-07 |
| FGF21* | Fibroblast growth factor 21 | Q9NSA1 | 2.101903 | 4.28E-06 |
| FGF21** | Fibroblast growth factor 21 | Q9NSA1 | 1.928575 | 7.79E-06 |
| IGFBP1 | Insulin-like growth factor-binding protein 1 | P08833 | 1.594791 | 7.98E-06 |
| t-PA (PLAT) | Tissue-type plasminogen activator | P00750 | 1.000429 | 9.52E-06 |
| IGFBP2 | Insulin-like growth factor-binding protein 2 | P18065 | 1.090845 | 1.4E-05 |
| CCL20 | C-C motif chemokine 20 | P78556 | 1.286329 | 1.46E-05 |
| CHI3L1 | Chitinase-3-like protein 1 | P36222 | 1.017746 | 0.000376 |
| CXCL10 | C-X-C motif chemokine 10 | P02778 | 1.091308 | 0.00046 |
| MMP1 | Interstitial collagenase | P03956 | 1.05074 | 0.003264 |
| PON3 | Serum paraoxonase/lactonase 3 | Q15166 | -1.08773 | 4.54E-09 |
| TNFSF12 (TWEAK) | TNF ligand superfamily member 12 | O43508 | -2.1196 | 0.000165 |
| PAPPA | Pappalysin-1 | Q13219 | -1.46461 | 0.004191 |
| CCL28 | C-C motif chemokine 28 | Q9NRJ3 | -1.76228 | 0.005024 |
| SPON1 | Spondin-1 | Q9HCB6 | -1.16843 | 0.01652 |
| **Biomarkers of high PVR in HF** (high PVR HF vs low PVR HF comparison) | | | | |
| CCL3* | C-C motif chemokine 3 (Macrophage inflammatory protein 1-alpha) | P10147 | 0.447285 | 0.029744 |
| CCL3** | C-C motif chemokine 3 (Macrophage inflammatory protein 1-alpha) | P10147 | 0.481298 | 0.029744 |
| PSP-D (SFTPD) | Pulmonary surfactant-associated protein D | P35247 | 0.718515 | 0.029744 |
| TNFRSF13B | TNF receptor superfamily member 13B | O14836 | 0.363009 | 0.035797 |

In brackets are newer abbreviations (from web https://olink.com/products/olink-target-96, in suppl 1).

For Uniprot ID see supplement 1. * in Olink panel CVII, ** in Olink panel Inflammation, *** in Olink panel CVIII
