## Supplementary material for "Transpulmonary proteome gradients identify pathways involved in the development of pulmonary vascular disease in heart failure": Suppl FIgure 1-3

### Slide 1
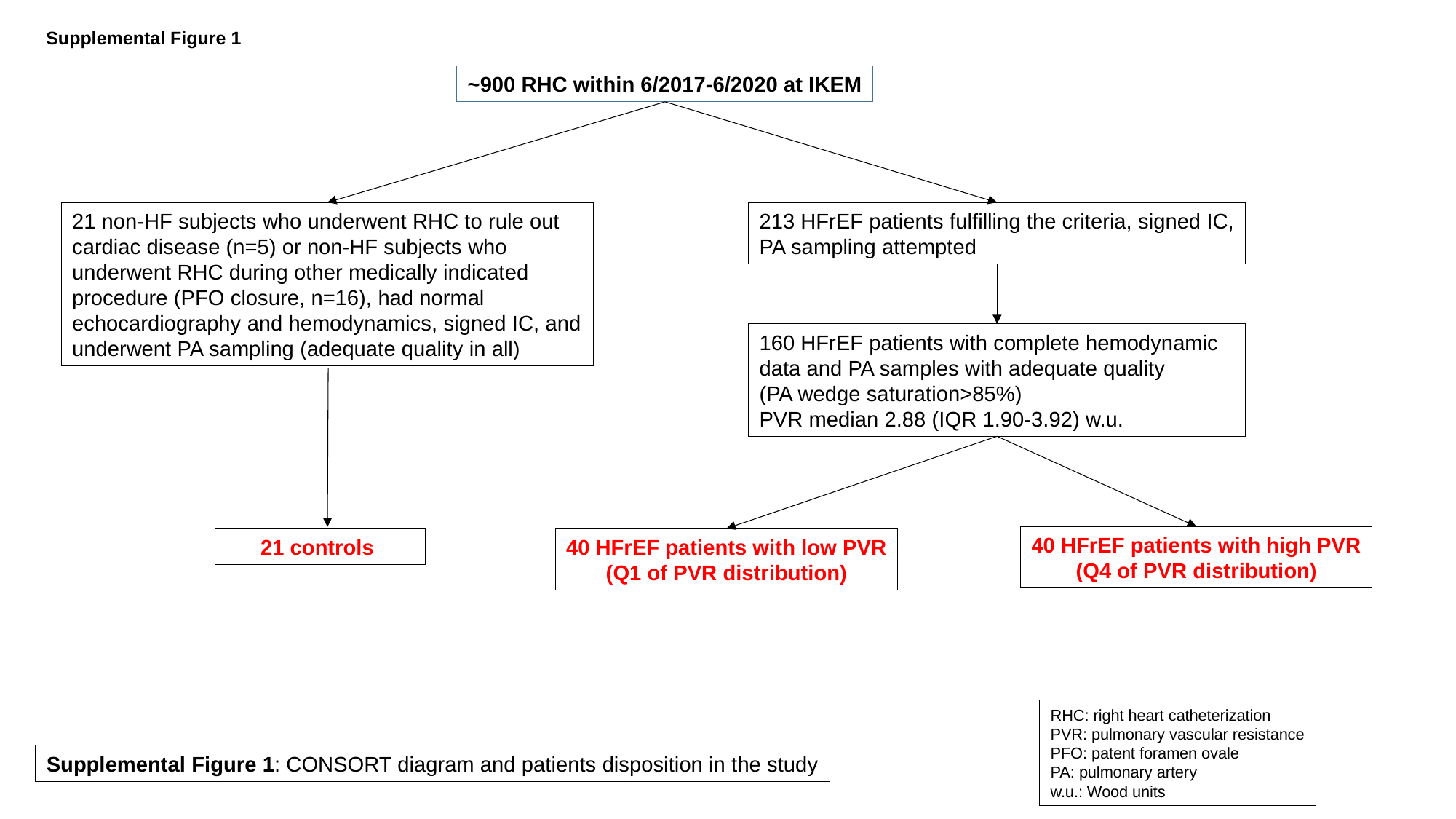

Supplemental Figure 1
~900 RHC within 6/2017-6/2020 at IKEM
21 non-HF subjects who underwent RHC to rule out cardiac disease (n=5) or non-HF subjects who underwent RHC during other medically indicated procedure (PFO closure, n=16), had normal echocardiography and hemodynamics, signed IC, and underwent PA sampling (adequate quality in all)
213 HFrEF patients fulfilling the criteria, signed IC,
PA sampling attempted
160 HFrEF patients with complete hemodynamic data and PA samples with adequate quality
(PA wedge saturation>85%)
PVR median 2.88 (IQR 1.90-3.92) w.u.
40 HFrEF patients with high PVR
(Q4 of PVR distribution)
21 controls
40 HFrEF patients with low PVR
(Q1 of PVR distribution)
RHC: right heart catheterization
PVR: pulmonary vascular resistance
PFO: patent foramen ovale
PA: pulmonary artery
w.u.: Wood units
Supplemental Figure 1: CONSORT diagram and patients disposition in the study

### Slide 2
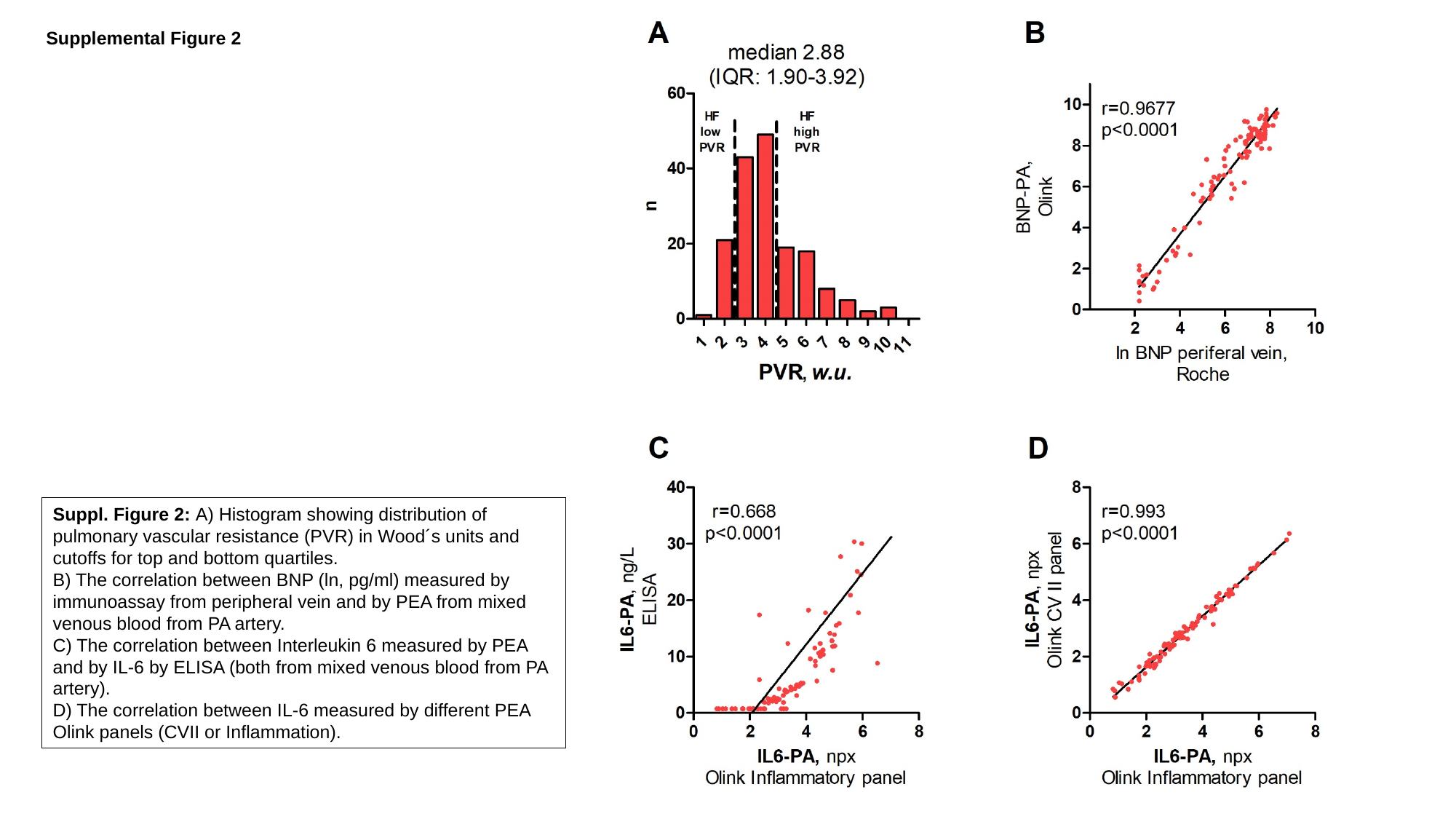

Supplemental Figure 2
Suppl. Figure 2: A) Histogram showing distribution of
pulmonary vascular resistance (PVR) in Wood´s units and cutoffs for top and bottom quartiles.
B) The correlation between BNP (ln, pg/ml) measured by immunoassay from peripheral vein and by PEA from mixed venous blood from PA artery.
C) The correlation between Interleukin 6 measured by PEA and by IL-6 by ELISA (both from mixed venous blood from PA artery).
D) The correlation between IL-6 measured by different PEA Olink panels (CVII or Inflammation).

### Slide 3
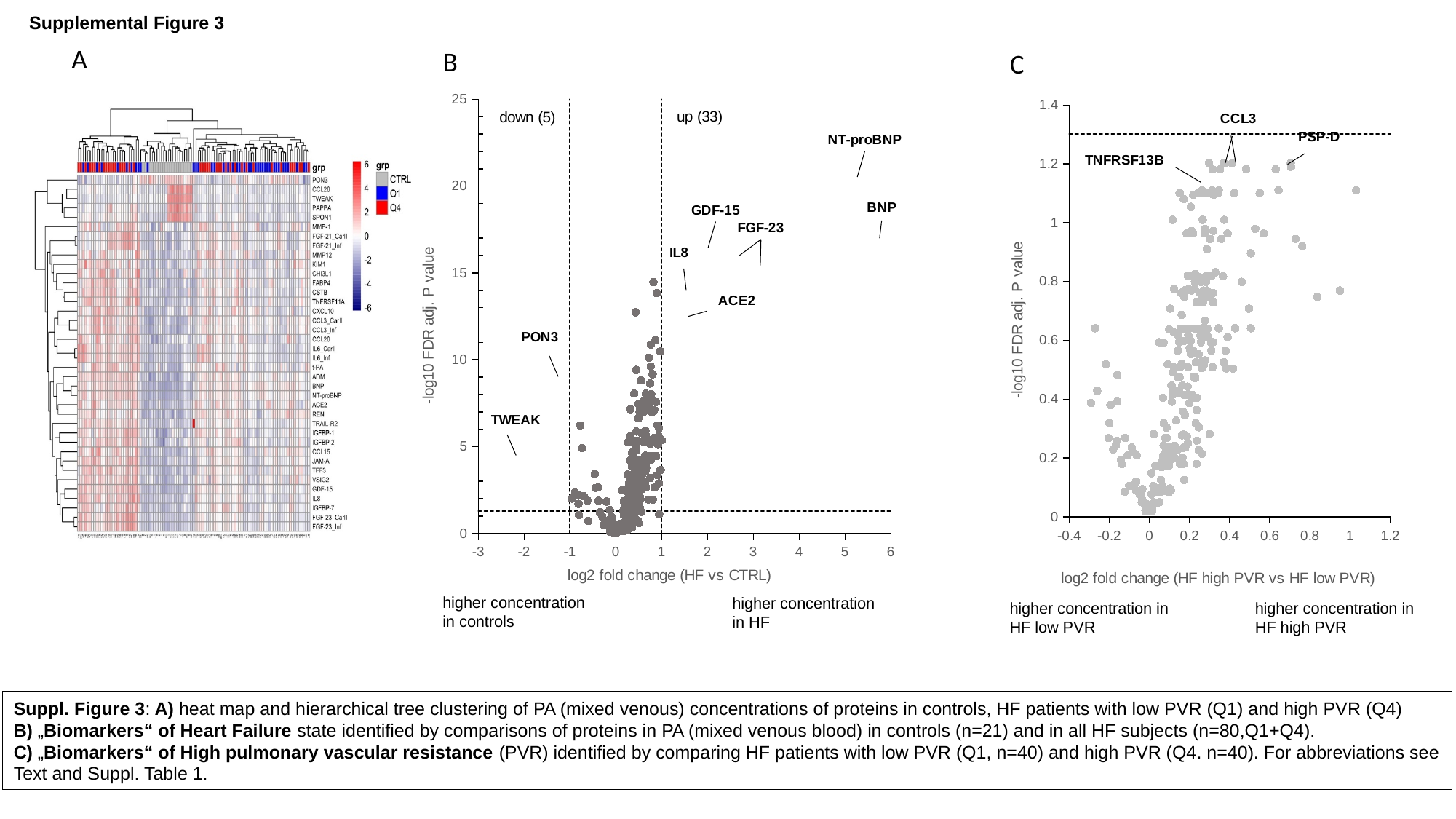

Supplemental Figure 3
A
B
C
#### Chart
| Category |
|---|
#### Chart
| Category |
|---|
higher concentration
in controls
higher concentration
in HF
higher concentration in
HF low PVR
higher concentration in
HF high PVR
Suppl. Figure 3: A) heat map and hierarchical tree clustering of PA (mixed venous) concentrations of proteins in controls, HF patients with low PVR (Q1) and high PVR (Q4)
B) „Biomarkers“ of Heart Failure state identified by comparisons of proteins in PA (mixed venous blood) in controls (n=21) and in all HF subjects (n=80,Q1+Q4).
C) „Biomarkers“ of High pulmonary vascular resistance (PVR) identified by comparing HF patients with low PVR (Q1, n=40) and high PVR (Q4. n=40). For abbreviations see
Text and Suppl. Table 1.

### Slide 4
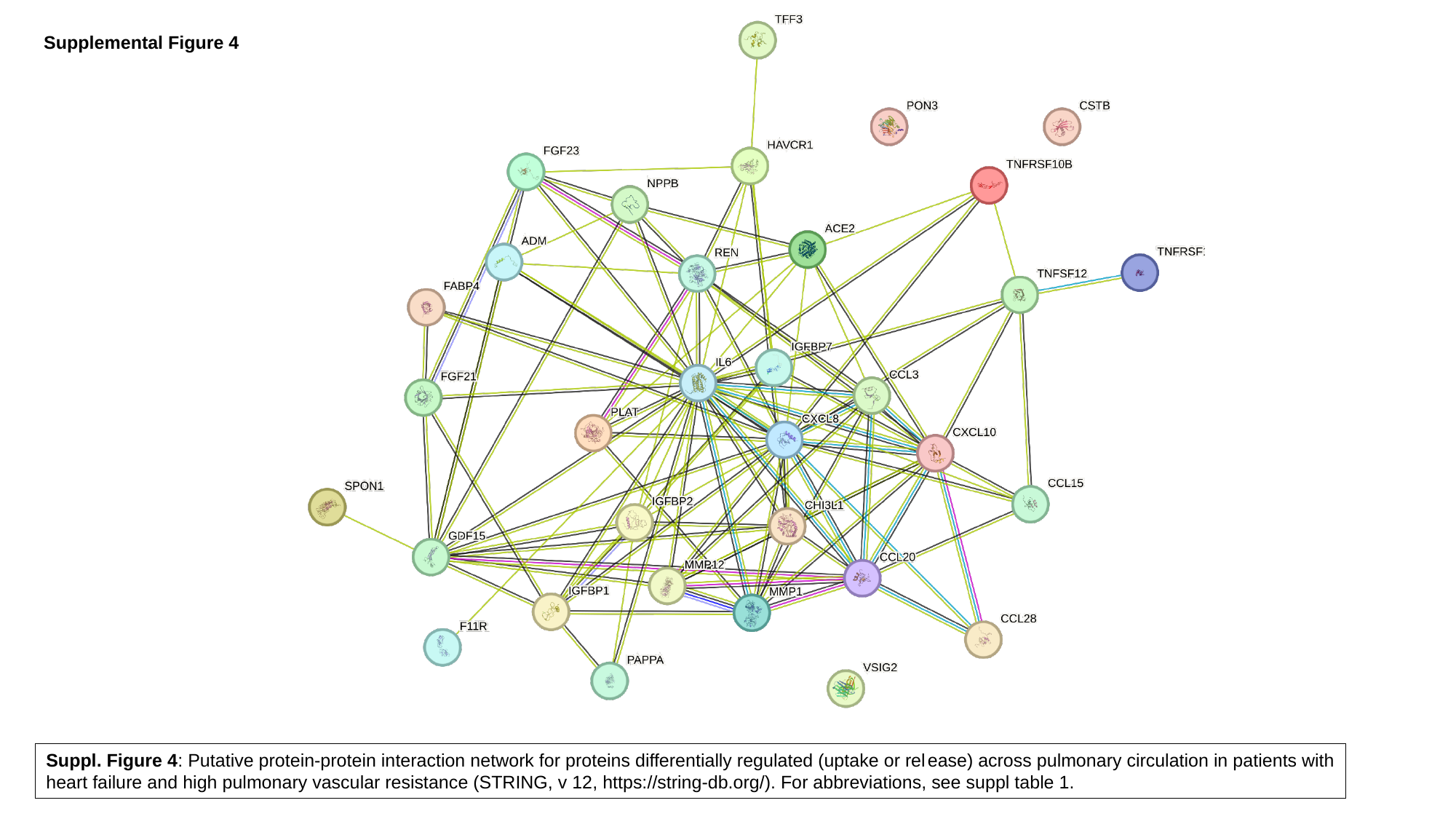

Supplemental Figure 4
Suppl. Figure 4: Putative protein-protein interaction network for proteins differentially regulated (uptake or release) across pulmonary circulation in patients with
heart failure and high pulmonary vascular resistance (STRING, v 12, https://string-db.org/). For abbreviations, see suppl table 1.
